# State-dependent non-identifiability of the reproduction number under adaptive behavior: an empirical characterization from COVID-19 mobility

**DOI:** 10.64898/2026.07.19.26358437

**Authors:** Fabio Sanchez

**Affiliations:** CIMPA–Escuela de Matemática, Universidad de Costa Rica

## Abstract

The basic reproduction number *R*_0_ confounds pathogen biology with adaptive human contact behavior. Earlier epidemiological–economic theory predicted a forward-looking behavioral contact response but could not test it in the absence of appropriate behavioral data. Using directly measured mobility as an observable proxy for contact, we (i) estimate the behavioral response function directly from data; (ii) show that the biology/behavior decomposition and hence the behavioral correction to *R*_0_ is not identified from an epidemic trajectory, the apparent constant-contact *R*_0_ being one endpoint of an observational-equivalence class that fits the factual curve identically yet diverges under counterfactual; and (iii) characterize that divergence (“what *R*_0_ deletes”) as *state-dependent*, unimodal in counterfactual severity and vanishing when behavior saturates. We then show that, across US jurisdictions, the correction is empirically bounded because risk-responsiveness and behavioral non-saturation are confounded (*r* = −0.57, *n* = 51): where behavior could compensate, it was already maximal, and where it was not maximal it did not respond. What *R*_0_ deletes is thus real and structurally characterizable yet empirically modest here, for reasons the framework itself supplies.

## 1 Introduction

The basic reproduction number *R*_0_ is the organizing scalar of mathematical epidemiology: the expected number of secondary infections produced by a typical case in a fully susceptible population, and the threshold whose crossing marks the onset of an epidemic [1]. Its authority rests on an implicit premise that the contacts through which transmission occurs are a fixed property of the population. That premise fails whenever people change their behavior in response to the epidemic itself. [2] made the point sharply: *R*_0_ is a reduced-form quantity that folds together the biology of the pathogen and the contact behavior of hosts, so that an estimate obtained during one behavioral regime need not describe transmission under another. In their words, *R*_0_ “represents past events that may not be informative of future events when behavior is adaptive.” The transmission rate that a constant-contact model attributes to biology is in fact the product of a biological susceptibility and a behavioral contact rate that is itself endogenous to prevalence.

Efforts to place behavior inside transmission models fall into three broad traditions. The first specifies incidence as a nonlinear function of the infected fraction, so that transmission saturates or declines as prevalence rises [3]; this is a reduced-form description of behavioral feedback that does not represent the decision generating it. The second adds behavioral compartments or agent-based rules: fear states, awareness classes, or individual heuristics [4], which requires the modeler to posit *a priori* how incentives map to behavior. The third, the epidemiological–economic tradition to which [2] belongs, derives contact as the solution of a forward-looking optimization in which individuals trade the utility of contact against the risk of infection [5]. Each tradition confronts the same obstacle: the behavioral object of interest, how contact actually responds to perceived risk, was not directly observable, so it had to be assumed. Reviews of the field have long identified this as the central empirical gap [6].

The COVID-19 pandemic removed that obstacle. Population mobility, measured continuously and at fine spatial resolution, provides a direct, if imperfect, observation of the very contact behavior that earlier models could only infer. A large body of work has since incorporated measured mobility into transmission models: to explain the timing and magnitude of transmission declines [7, 8], to attribute reductions in the reproduction number to mobility change [9], and to resolve fine-grained spatial and demographic heterogeneity in spread [10]. Our question is orthogonal to all of these and, to our knowledge, unasked in them: not how well mobility *explains* the observed epidemic, but what the epidemic curve can *identify* about the biology/behavior split, and how behavior would re-respond off the observed path. Almost universally in this literature mobility enters as *exogenous forcing* : the contact time series is treated as a given input, and the model reproduces the observed epidemic by fitting biology to it. This is observationally adequate; it fits, but it discards precisely the feature that made behavior consequential in the first place. Exogenous forcing assumes that, under a counterfactual (a more transmissible variant, an earlier intervention, a different policy), behavior would have followed the same path it in fact took. Endogenous behavior would not: it would have re-responded to the altered epidemic. The exogenous-forcing literature, therefore, cannot address the counterfactual questions for which behavior matters, and, to our knowledge, has neither asked what is actually identified once behavior is allowed to be endogenous nor estimated the response function itself.

We take up both questions. First, we estimate the behavioral response function contact as a function of lagged perceived risk directly from data, providing an empirical counterpart to the forward-looking response posited by [2]. Second, we show that the decomposition of the observed transmission into a biological rate and a behavioral response is *not identified* from an epidemic trajectory: a one-parameter family of models, differing in how much of the observed contact change is treated as endogenous, reproduces the factual epidemic identically and yields the same apparent *R*_0_ yet gives different answers under counterfactual. The constant-contact *R*_0_ is the extreme, fully-exogenous member of this equivalence class. Third, we characterize the counterfactual divergence of the “deleted” behavioral content and find it *state-dependent* : unimodal in counterfactual severity, maximal at intermediate severity, and vanishing both when the epidemic barely occurs and when behavior is already saturated. Finally, using a cross-jurisdiction panel, we show why the correction is empirically small in practice: the two conditions that would make it large strong risk-responsiveness and behavioral non-saturation are negatively correlated across US states and never co-occur.

Our conclusion is deliberately double-edged. The behavioral content that *R*_0_ omits is real, and we give it a precise structural characterization rather than a vague warning. But in the COVID-19 setting that content is empirically modest, and the same framework that identifies it also explains why. We regard honest delimitation of an effect as more useful than its inflation.

## 2 Data

### Cases, hospitalizations, deaths

New York City daily counts by event date (case, hospitalization, death), NYC DOHMH [11]. US state-level cases and deaths, [12]. Both indexed by event date.

### Mobility (contact proxy)

Google COVID-19 Community Mobility Reports [13], retrieved from the archived mirror [14]. Reports give percent change from the Jan 3–Feb 6 2020 median, so the normalized contact index *C*(*t*) = 1 + pct(*t*)*/*100 satisfies *C* = 1 at the pre-epidemic baseline by construction. NYC series are population-weighted over the five boroughs (New York, Kings, Queens, Bronx, Richmond counties); state series use the state-level (`county=“Total”`) records. Primary category: transit stations for NYC, retail & recreation for the cross-state panel (transit is uninformative in low-transit states).

### Fixed epidemiological inputs

(assumptions; sensitivity in §4.7): removal rate *v* = 1*/*6.5 d^−1^ [15]; generation interval ~ Γ(mean 5.2, sd 3.9) [16]; infection*→* hospitalization delay ~ Γ(11, 5) and infection*→* death delay ~ Γ(18, 8) [17].

## 3 Model and methods

### 3.1 Behavior-explicit transmission with measured contact

With susceptible, infectious, and removed fractions *S* + *I* + *Z* = 1 and *C*(*t*) an observed input,

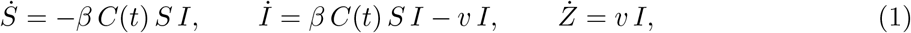

so that *Z* (recovered or removed, including deaths) is determined by *Z* = 1 − *S* − *I* and does not feed back into transmission. Only the product *βC*(*t*) enters the dynamics. Fixing *C*(baseline) = 1 identifies *β* as transmission per unit baseline contact. The deep utility parameters that generate *C*(*t*) in [2] are *not* identified here; instead, we replace the structural behavior model with its measured output. Mobility is a proxy for contact, not contact itself, so *β* absorbs the (possibly nonlinear) mobility-to-contact map.

### 3.2 Object A: apparent *R*_0_

Under the analyst’s constant-contact assumption, the early exponential growth rate 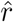maps to 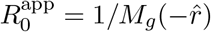, with *M*_*g*_ the generation-interval moment-generating function [18] (Appendix A); for the SIR special case 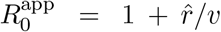. We estimate 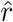by ordinary least squares of log(hospitalizations by date) on time over 8–22 March 2020 (the log-linear window; *n* = 15 days), giving 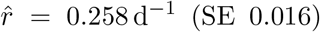. The estimate is moderately window-sensitive: across eight start/end choices spanning 5–26 March, 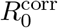 ranges over [2.36, 3.14] about the central value 2.71 itself a manifestation of the practical non-identifiability of any scalar reduction (§4.4), not of the composite *βC*(*t*).

### 3.3 Object B: growth-phase identification of *β*

During exponential growth (*S ≈* 1), d log *I/*d*t* = *βC*(*t*) − *v*. We estimate *β* from the observed growth rate and the mean contact over the *infection* window that generated the observed (delayed) event window: 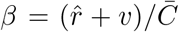. This growth-phase estimand is well-specified precisely where the constant-contact analyst operates and where mobility best proxies contact; it avoids the post-peak regime where the well-mixed model is misspecified.

### 3.4 The behavioral response function *R*

People reduce contact in response to *perceived* risk (reported deaths), not latent prevalence. We estimate

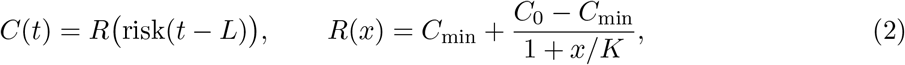

with risk = reported deaths (7-day average) and behavioral lag *L*, by nonlinear least squares. This is the empirical analog of the forward-looking optimizing response of [2]: we estimate the realized input–output map rather than the utility primitives that generate it.

### 3.5 Non-identifiability and the equivalence class

An *exogenous* model integrates (1) with *C*(*t*) frozen at its factual path *C*_fac_; an *endogenous* model sets *C*(*t*) = *R*(risk[*I*](*t* − *L*)), so that contact tracks the model’s own delayed deaths. On the factual path the two coincide by construction, *C*_fac_(*t*) = *R*(risk_fac_(*t* − *L*)) up to the fit residual of (2); they are therefore observationally equivalent in-sample and differ only in how contact would *re-respond* to a perturbation. To index that difference, introduce the counterfactual contact under a transmissibility perturbation *f*,

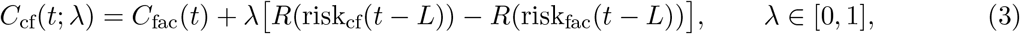

where risk_cf_ is the risk path generated under the perturbed dynamics and risk_fac_ the factual one.

#### Status of the construction

Equation (3) is a one-parameter parameterization of *partial endogeneity*, not a linearized approximation. It is built to (i) anchor the exogenous endpoint exactly on the observed contact at *λ* = 0, *C*_cf_ = *C*_fac_, so the factual fit is reproduced exactly, independent of the quality of the response fit (2) and (ii) recover full endogeneity at the other endpoint at *λ* = 1, contact responds fully to the counterfactual risk, *C*_cf_ = *C*_fac_ + *R*(risk_cf_) − *R*(risk_fac_). Writing the response gap Δ*R*(*t*) = *R*(risk_cf_(*t* − *L*)) − *R*(risk_fac_(*t* − *L*)), the family is the line *C*_fac_ + *λ* Δ*R*. Three readings therefore coincide for this two-endpoint construction: (a) modulo the factual residual it is the *convex combination* (1 − *λ*)*R*(risk_fac_) + *λR*(risk_cf_); (b) its endpoint derivative *∂*_*λ*_*C*_cf_|_*λ*=0_ = Δ*R* is the *first-order* sensitivity of contact to the perturbation; and (c) *λ* is the *fraction of endogeneity* activated under counterfactual. We adopt the linear form for its transparency, but the non-identifiability below does not rely on it (Prop. 2).

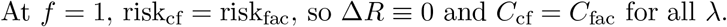

##### Proposition 1

(Observational equivalence). *For every λ ∈* [0, 1], *the model* (1) *with contact* (3) *and f* = 1 *produces an identical factual trajectory and hence identical apparent R*_0_. *λ is not identified from* (*y*(*t*), *C*(*t*)) *on the observation window. Counterfactuals (f* ≠ 1*) that perturb the risk path are functions of the non-identified λ; λ* = 0 *(fully exogenous) is the apparent-R*_0_ *endpoint*.

*Proof*. At *f* = 1 the two risk paths coincide, risk_cf_(*·*) = risk_fac_(*·*), so the bracketed term in (3) vanishes identically in *t* and *C*_cf_(*t*) *≡ C*_fac_(*t*) independently of *λ*. Consequently, the solution of (1) is the same for every *λ*, as is every functional of it, in particular the incidence *y*(*t*), the growth rate 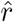, and 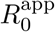. Thus no feature of (*y*(*t*), *C*(*t*)) depends on *λ*, which is therefore not identified. For *f* ≠ 1 the perturbation alters risk_cf_, the bracketed term is generically nonzero, and *C*_cf_ hence the counterfactual trajectory depends linearly on *λ*; the exogenous endpoint *λ* = 0 freezes contact at its factual path and coincides with the constant-contact 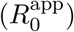 prediction. □

We verify the equivalence numerically to integrator tolerance (Fig. 2, right): the *λ*-family is graphically indistinguishable on the factual curve and fans out under counterfactual. Because at *f* = 1 the contact input *C*_cf_ is identical across *λ* by construction, the integrated trajectories are identical to machine precision (maximum absolute incidence difference between *λ* = 0 and *λ* = 1 is 0); the equivalence is exact, not approximate. Figure 1 makes the geometry explicit: at *f* = 1 the entire *λ*-family collapses onto the single observed curve, while at a counterfactual *f <* 1 it fans into the identified set whose width is “what *R*_0_ deletes.”

**Figure 1:**
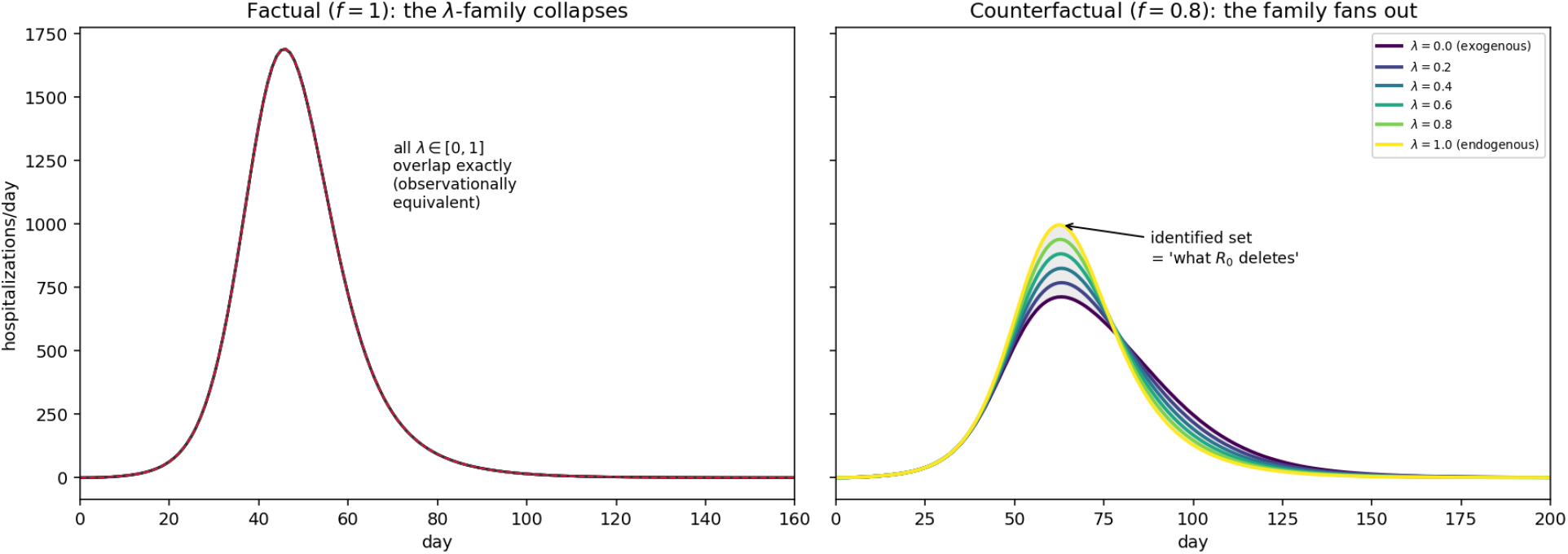
The observational-equivalence class, computed from the fitted model. Both panels show daily hospitalizations (model output, per day) against time in days; curves are the *λ*-family of (3) for *λ* = 0, 0.2, …, 1. **Left:** at *f* = 1 the entire family produces the identical factual curve the data cannot distinguish the endogenous fraction *λ*. **Right:** under a counterfactual transmissibility *f* = 0.8 the family fans out between the exogenous endpoint (*λ* = 0, contact frozen at its factual path) and the fully endogenous endpoint (*λ* = 1, contact re-responding to the altered risk). The shaded width the vertical span of the family is the identified set, “what *R*_0_ deletes.”

**Figure 2:**
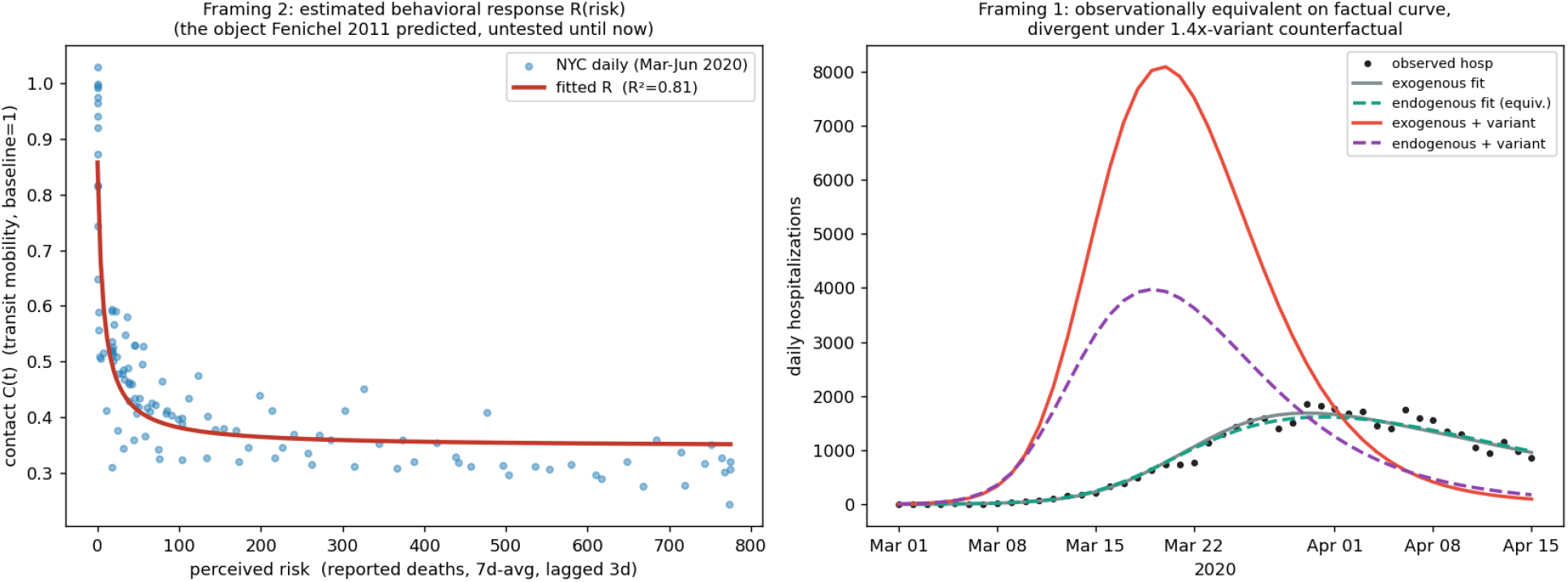
**Left:** estimated behavioral response *R* (Eq. 2), *R*^2^ = 0.81. **Right:** observational equivalence: exogenous and endogenous models coincide on the factual curve and diverge under a 1.4*×* transmissibility counterfactual.

The construction (3) is one chart of the identified set; the non-identifiability is a property of the equivalence at *f* = 1, not of the linear parameterization, as the following makes precise.

##### Proposition 2

(Specification-invariance of the non-identifiability). *Let* {Φ_*θ*_}_*θ*∈Θ_ *be any family of counterfactual contact rules* 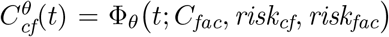 *satisfying the factual-consistency condition*

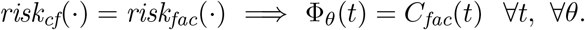

*Then at f* = 1 *the trajectory is independent of θ, and θ is not identified from* (*y*(*t*), *C*(*t*)) *on the observation window*.

*Proof*. At *f* = 1 the perturbation is absent, so risk_cf_ = risk_fac_ and, by hypothesis, Φ_*θ*_(*t*) = *C*_fac_(*t*) for every *θ*. The right-hand side of (1) is then *θ*-independent, hence so is its solution and every functional thereof. □

##### Remark 1

(Robustness to alternative specifications of partial endogeneity). *The scalar-λ family (3)is the minimal chart satisfying the hypothesis of Prop. 2. Two natural enrichments leave the conclusion intact and, if anything, strengthen it*. (a) Nonlinear interpolation: *replacing λ by g*(*λ*) *with g*(0) = 0, *g*(1) = 1 *traces the same contact paths under a smooth reparameterization, so the identified set is unchanged*. (b) State-dependent endogeneity: *allowing λ* = *λ*(*t*) *or λ* = *λ*(*I*) *with range* [0, 1] *replaces the line segment by its pointwise convex hull an infinite-dimensional identified set that* contains *the scalar one. In every case* Δ*R ≡* 0 *at f* = 1, *so the whole family collapses to the factual trajectory in-sample. The scalar λ therefore yields a* conservative *identified set: the counterfactual spans reported in §4*.*6 and §4*.*7 are lower bounds on those admitted by richer partial-endogeneity models*.

### 3.6 The regime map

For a grid of counterfactual transmissibility factors *f* we integrate (1)+(3) and record the peak and final size for *λ ∈* [0, 1]. The width of the resulting identified set the span over *λ* is the operational measure of “what *R*_0_ deletes.”

#### Integration

The system is integrated deterministically by a fixed-step explicit (Euler) scheme sub-stepped 8*×* per day (Δ*t* = 1*/*8 d), with contact held at its daily value within sub-steps; delays are applied as discrete convolutions with the gamma kernels of §2 (infection*→* death for the risk signal, infection*→* event for the observed series). The horizon is 400 d, which is past epidemic completion at every *f* considered: the change in cumulative attack rate over the final 20 d is *<* 10^−5^ even for the slowest (*f* = 0.6) epidemic, so final size is read at the terminal time without a separate stopping rule. The factual reference (ifr, *I*_0_, *ρ*) is calibrated once by nonlinear least squares to the March 1–April 15 2020 hospitalization series; peak ratios are reported relative to this factual peak, so the reporting factor *ρ* cancels.

#### Uncertainty

We propagate parameter uncertainty into the span by a bootstrap: for each of 150 replicates we resample the response-fit data (2) and refit (*C*_0_, *C*_min_, *K*), and draw the growth rate 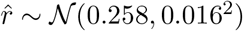 (log-linear regression SE) so that 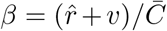; each replicate is recalibrated and rerun over the full *f*-grid, yielding a distribution of the span at every *f* (median and 5th–95th percentiles in Fig. 5).

### 3.7 Cross-jurisdiction panel

For each US state and the District of Columbia we form two summaries from 2020 data. *Risk-responsiveness* is the coefficient of determination *R*^2^ (floored at 0) of the state-level saturating fit (2), with contact *C* = 1 + pct*/*100 from the retail & recreation series and perceived risk the 7-day moving average of daily death increments (clipped at zero) from [12], at behavioral lag *L* = 3 d, over 1 March–31 December 2020. It measures how tightly contact tracked perceived risk. *Saturation* is summarized by min *C*, the minimum of the 7-day centered moving average of *C* over 2020; a lower min *C* means contact fell further, i.e. less remaining headroom. We test whether high responsiveness and high headroom (non-saturation) co-occur the joint condition under which the deletion of §3.6 would be large.

#### Low-incidence states

A state enters only if the fit has at least 80 valid daily observations and a strictly positive peak risk; this drops states whose death series is too sparse to identify a response. Because a weak death signal can still yield an unstable *R*^2^, we additionally report all associations after excluding the bottom quartile of states by peak 7-day deaths (§4.6).

## 4 Results

### 4.1 The behavioral response function is estimable and tight

In NYC, the response (2) fits with *R*^2^ = 0.81 (0.77–0.93 across risk proxies and lags; §4.2) using transit mobility, perceived risk = reported deaths (7-day average), and a behavioral lag *L* = 3 d: contact declines from *C*_0_ = 0.857 at zero perceived risk to a floor *C*_min_ = 0.347, with half-saturation *K* = 7.3 deaths/day (Fig. 2, left). The lag is chosen for identification rather than fit so that contact responds to *already-observed* risk and is conservative: the fit improves monotonically as *L →* 0 (*R*^2^ = 0.93 at *L* = 0; §4.2). The response is empirically consistent with the qualitative form of the forward-looking contact response posited by [2]: contact falls steeply with initial perceived risk and saturates at a positive floor, consistent with a bounded willingness to forgo contact. We stress the qualitative nature of this correspondence: [2] derives no closed-form *R*(*x*), so the comparison is of shape (monotone decline, saturation, positive floor, diminishing marginal response), not of functional form.

### 4.2 Robustness of the response function

We probed the fit along three axes (Fig. 3); details follow.

**Figure 3:**
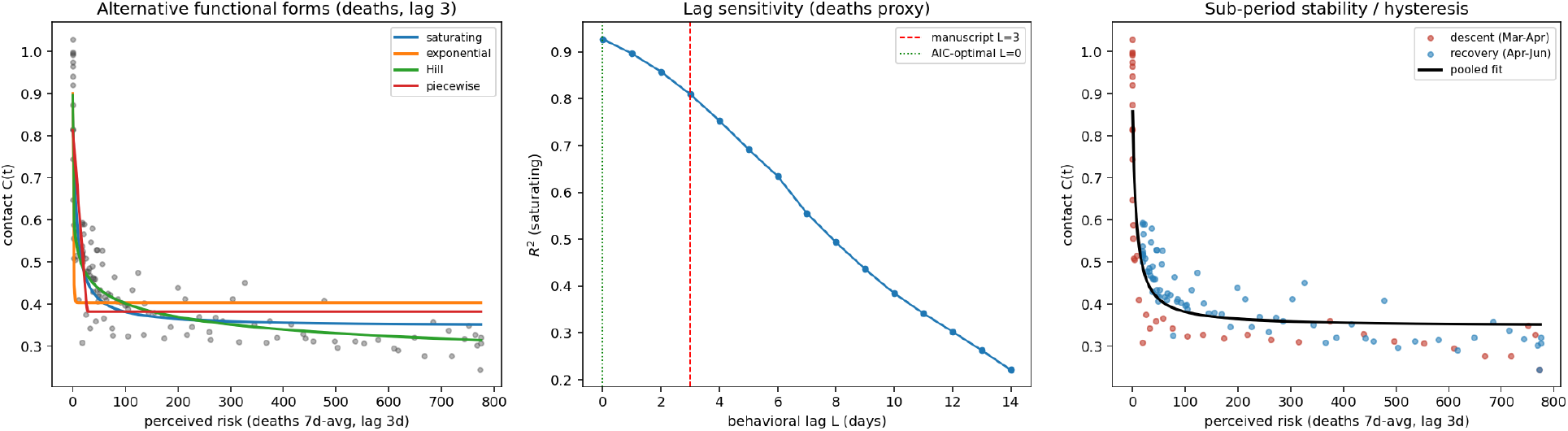
Robustness of the response function *R* (§4.2). **Left:** alternative functional forms on the deaths proxy (*L* = 3); exponential and clipped-linear reach the floor too abruptly. **Center:** *R*^2^ versus behavioral lag *L*; the fit is best at *L* = 0 and declines monotonically, with *L* = 3 retained on identification grounds. **Right:** descent (Mar–Apr) and recovery (Apr–Jun) phases lie on a common curve (no systematic hysteresis); the recovery phase is noisier.

#### Functional form

Fitting four forms to the same data (deaths, *L* = 3; Table 1), the saturating form (2) is preferred by BIC among the parsimonious (three-parameter) candidates; the exponential and clipped-linear forms fit worse because they reach the floor too abruptly. A four-parameter Hill form attains a higher *R*^2^ (0.88) but only through a boundary solution (steepness *h* driven to its lower bound and the floor to 0.16), which we read as over-flexible rather than as evidence of a distinct shape; on the cases proxy the Hill exponent collapses to *h ≈* 1, recovering (2). We therefore retain (2) as a parsimonious description whose qualitative features are form-robust.

**Table 1:** Alternative functional forms for *R* (deaths proxy, *L* = 3, common sample). Among three-parameter forms the saturating form (2) is preferred; the Hill gain rests on a parameter-boundary solution (§4.2).

| functional form | $k$ | $R^2$ | AIC | BIC |
| --- | --- | --- | --- | --- |
| saturating (2) | 3 | 0.810 | −601.9 | −593.6 |
| exponential decay | 3 | 0.762 | −575.7 | −567.3 |
| clipped-linear (piecewise) | 3 | 0.710 | −551.9 | −543.6 |
| Hill (boundary soln.) | 4 | 0.878 | −653.0 | −641.9 |

#### Risk proxy and lag

All three proxies deaths, reported cases, and hospitalizations yield the same saturating shape with comparable fit (*R*^2^ in [0.77, 0.94]), each at its own information-criterion-optimal lag (deaths *L* = 0, cases *L* = 2, hospitalizations *L* = 5). By AIC the cases proxy is marginally best, but early-2020 case counts are confounded by testing expansion (the same artifact that inflates the case-based *R*_0_ in §4.4); we therefore use deaths as the mechanistically cleaner perceived-risk signal. The shape and floor are unchanged by the choice.

#### Sub-period stability

Splitting at the mobility trough (13 Apr 2020) into a descent and a recovery phase, a single response curve describes both adequately (a two-curve fit does not improve BIC), with no systematic hysteresis loop; the recovery phase is noisier (*R*^2^ = 0.78 vs 0.97 on descent), consistent with behavior partially decoupling from risk as reopening proceeds. The implied elasticity of contact with respect to risk is negative and declines in magnitude from −0.27 at low risk to −0.07 at high risk bounded, saturating avoidance, matching the qualitative Fenichel prediction.

### 4.3 Apparent *R*_0_ and its growth-phase correction

On the clean log-linear window (hospitalizations, 8–22 Mar 2020), 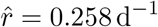, giving 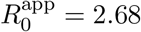 (SIR) / 2.72 (GI-MGF; 95% CI [2.48, 2.97]). The choice of proxy matters: reported cases inflate the estimate 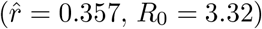 through test ramp-up, and deaths deflate it 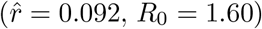 through sparse early counts; hospitalizations by date of hospitalization are the defensible lead.

Growth-phase identification (§3.3) gives mean contact 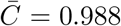 over the generating infection window (26 Feb–11 Mar), hence *β* = 0.417 and 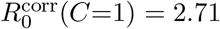. That this nearly equals 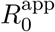is expected: early infections occurred at near-baseline contact, so the scalar bias at baseline is small. The often-stated intuition that behavior badly biases the *scalar R*_0_ is thus not supported here; the behavioral content lies elsewhere, in the counterfactual.

### 4.4 The decomposition is non-identified

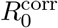 is stable to the fixed inputs (*v*, GI, delay) (2.50–2.66 over the sensitivity grid, §4.7) but *not* to specification: full-window fits including the misspecified post-peak plateau return 3.4–3.6. This specification-dependence is the empirical signature of non-identifiability (Prop. 1): the behavior-resolved composite *βC*(*t*) is invariant while the scalar reductions of it are assumption-dependent. Fig. 2 (right) shows the *λ*-family fitting the factual curve identically and diverging under a 1.4*×* transmissibility counterfactual; the exogenous (*λ* = 0) and endogenous (*λ* = 1) endpoints bracket a range of outcomes the data cannot distinguish.

### 4.5 “What *R*_0_ deletes” is state-dependent and unimodal

The identified-set span is *unimodal* in counterfactual severity (Fig. 4, Fig. 5): near zero for *f ≤* 0.6 (epidemic barely occurs) and for *f >* 1 (behavior already at floor: no compensation available), peaking at *f ≈* 0.8, where the deleted content reaches 0.18 on the peak ratio (bootstrap median; 90% band [0.15, 0.22]) as behavior rides the steep part of *R*. The unimodal shape is preserved under uncertainty propagation: the band at the *f* = 0.8 maximum is well separated from the near-zero span at *f* = 1. Final-size spans are far smaller median *≤* 2.5 % for *f ≥* 0.7, rising to *≈* 3 % (median; 95th percentile *≈* 6 %) only at the near-threshold value *f* = 0.6, where final size is intrinsically sensitive. The behavioral content that *R*_0_ deletes is thus real but bounded, largest at intermediate severity, and this is the substantive asymmetry almost entirely a matter of the *peak*, not the *attack rate*: adaptive behavior reshapes the height and timing of the surge (a peak-ratio swing of up to ~ 20%) while leaving the eventual cumulative incidence nearly fixed (a few percent). For planning this separates cleanly: the unidentified behavioral response matters for surge-capacity questions (peak hospital and ICU load, and *when* they arrive), but is largely irrelevant to the final attack rate, which is pinned by the trajectory almost independently of *λ*.

**Figure 4:**
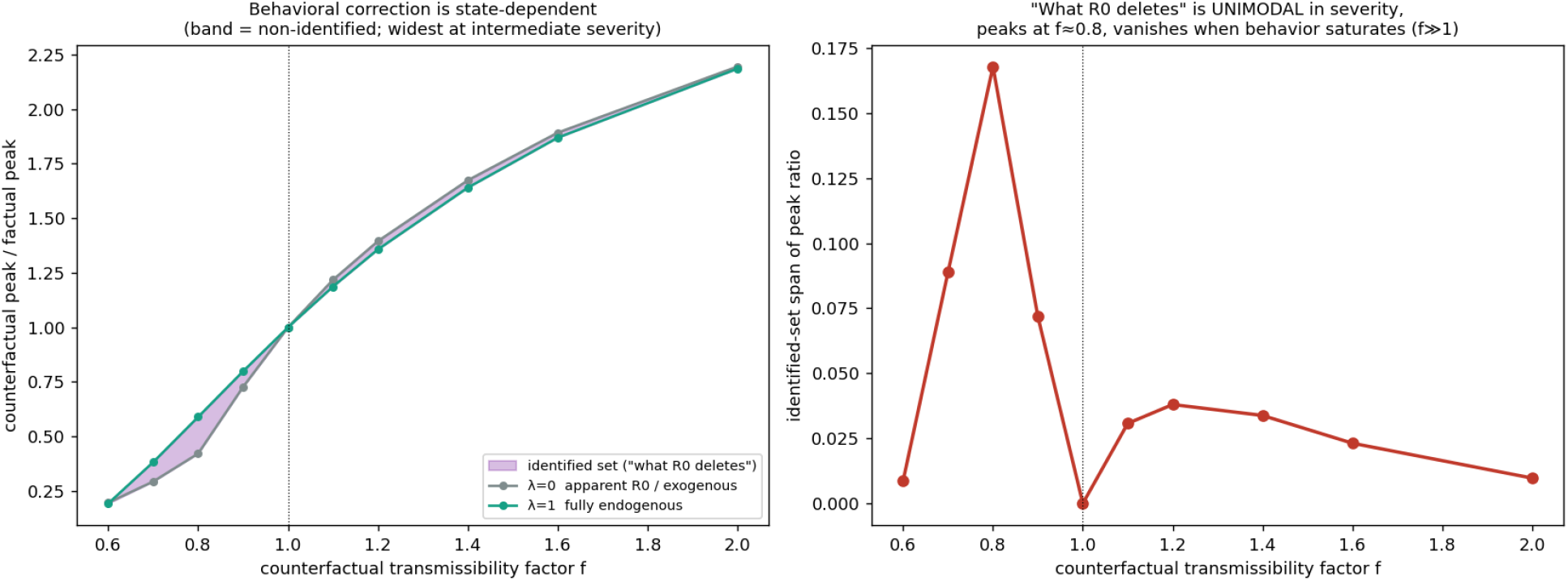
Identified-set span (“what *R*_0_ deletes”) is unimodal in counterfactual transmissibility *f* : near zero under saturation (*f >* 1) and when the epidemic barely occurs (*f ≤* 0.6), maximal at *f ≈* 0.8.

**Figure 5:**
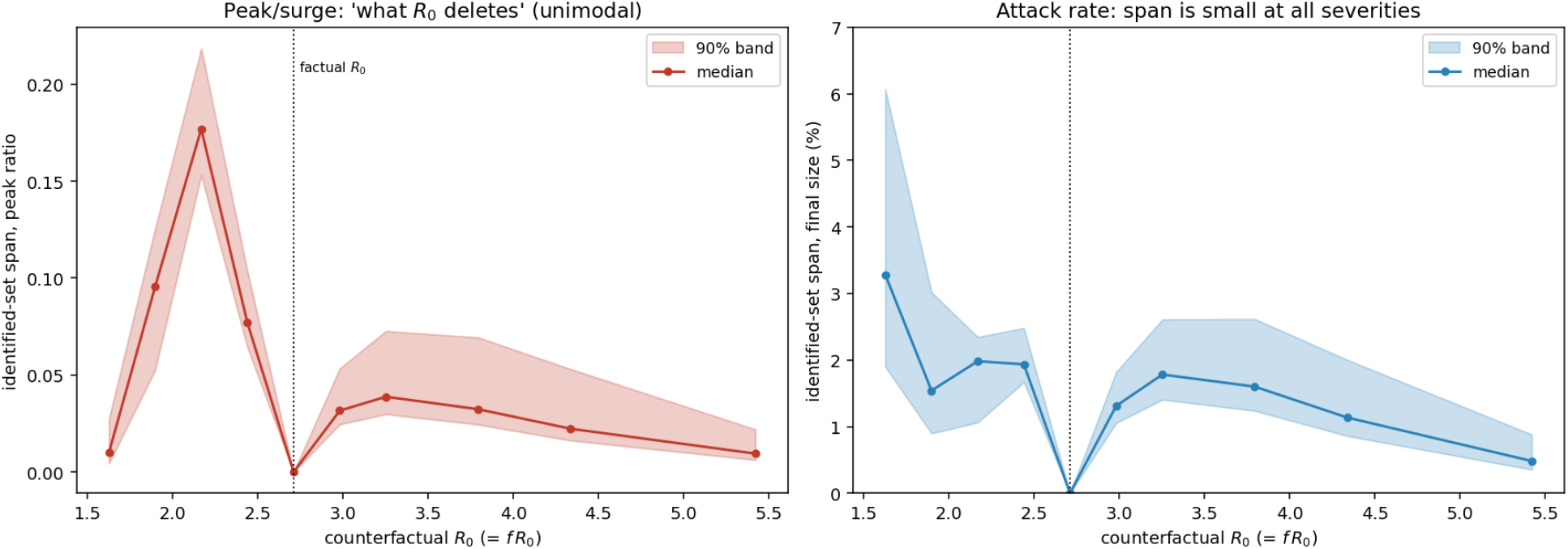
Uncertainty in the identified-set span. The span is the width of the *λ ∈* [0, 1] family at each counterfactual the peak-ratio range (left) and the final-size range in percentage points (right) the same quantity plotted in Fig. 4. Bands are from a bootstrap of *B* = 150 replicates that resample the response-fit data and draw the growth rate 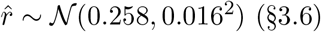; the line is the median and the shaded region the 5th–95th percentile (90%) band. **Left:** the peak-ratio span is unimodal and the shape survives propagation, peaking near counterfactual *R*_0_ *≈* 2.2 (*f* = 0.8). **Right:** the final-size span is an order of magnitude smaller (note the axis), ≲ 2.5 % except near the epidemic threshold. Behavior reshapes the surge, not the attack rate.

### 4.6 Responsiveness and non-saturation are confounded across states

Across *n* = 51 US jurisdictions, minimum mobility (saturation) ranges from 0.24 (DC) to 0.77 (Wyoming): strong cross-sectional variation. But risk-responsiveness (*R*^2^ of (2)) and non-saturation are negatively associated (*r* = −0.57, *p* = 1.3 *×* 10^−5^; Fig. 6). Strongly responsive states (*R*^2^ *>* 0.4; *n* = 2) all cratered mobility (min *C ∈* [0.34, 0.44]); unsaturated states (min *C >* 0.55; *n* = 26) show weak response (*R*^2^ *∈* [0.00, 0.27]). The “large-deletion” quadrant responsive *and* unsaturated is empty. Where behavior could have compensated, it was already maximal; where it was not maximal it did not respond. This explains why the behavioral correction, though real, is empirically modest: the conditions that would make it large do not co-occur in this setting.

**Figure 6:**
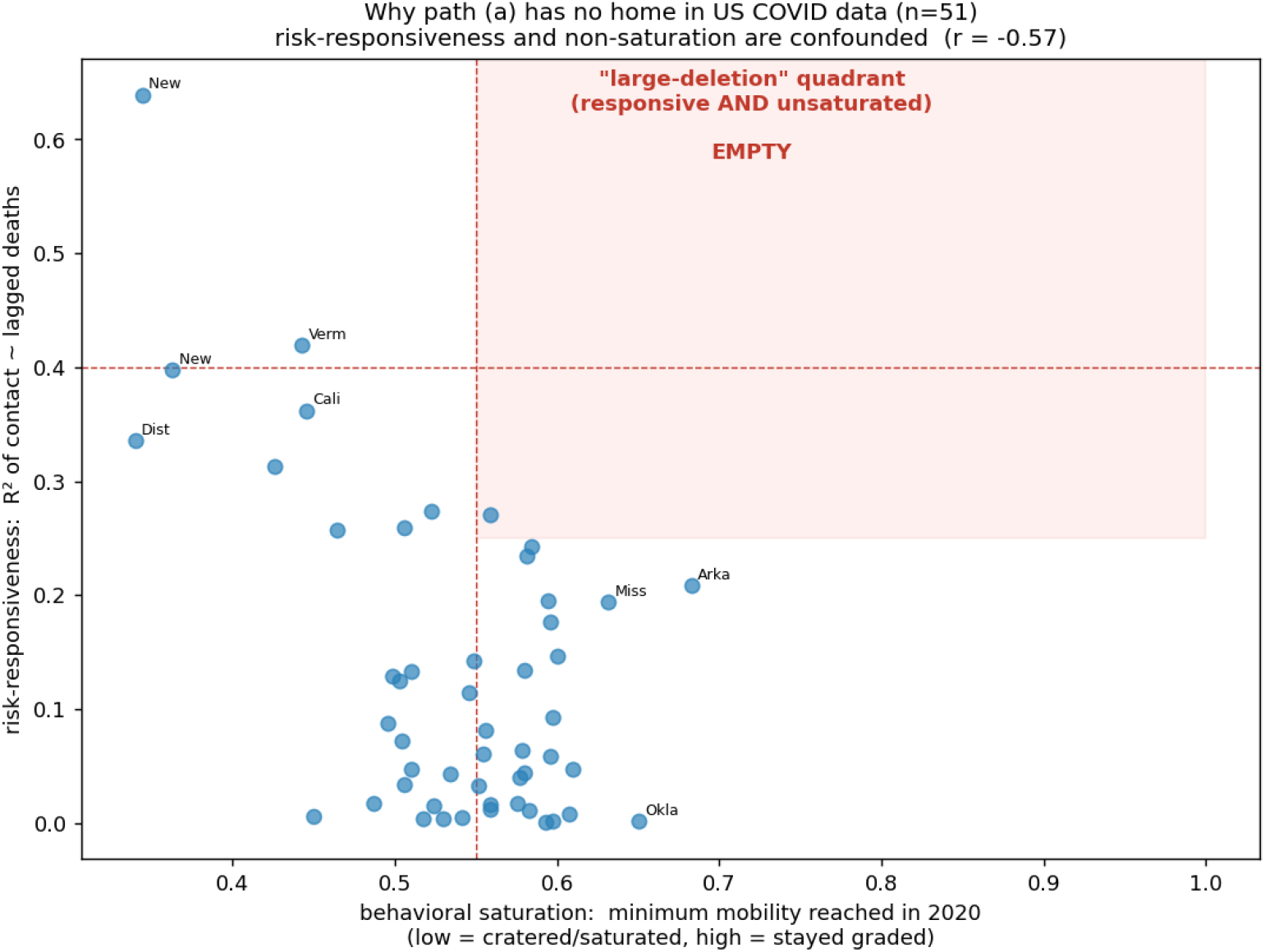
Risk-responsiveness versus non-saturation across the *n* = 51 US states and DC. Horizontal axis: per-state response-fit *R*^2^ (how tightly contact tracked perceived risk, §3.7); vertical axis: min *C*, the deepest 7-day-smoothed mobility trough in 2020 (higher = less saturated, more headroom). The two are negatively associated (Pearson *r* = −0.57, Spearman *ρ* = −0.32); the large-deletion quadrant (high *R*^2^ *and* high min *C*, upper right) is empty and remains so under the robustness checks of §4.6.

The association is robust but the Pearson coefficient overstates its rank strength, which we report to avoid overclaiming. The rank correlations are weaker though still significant (Spearman *ρ* = −0.32, *p* = 0.024; Kendall *τ* = −0.21, *p* = 0.028), so the relationship is moderate rather than strong. It is not driven by any single state (leave-one-out Pearson *r ∈* [−0.62, −0.47]), not inflated by outliers (removing the one state with |*z*| *>* 2.5, Arkansas, *strengthens* it to −0.62), and robust in sign under robust regression (Theil–Sen slope −0.21, 95% CI [−0.37, −0.03]). Excluding the bottom quartile of states by peak deaths where the response fit is least reliable leaves it unchanged (*n* = 38; Pearson −0.59, Spearman −0.37). Crucially for the paper’s claim, the empty large-deletion quadrant is invariant: it remains empty among high-incidence states and across the threshold choices (*R*^2^ *>* 0.3–0.5, min *C >* 0.5–0.6) we examined.

### 4.7 Sensitivity analysis

Over the grid *v* ∈ {1/7.5, 1/6.5, 1/5.5} d^−1^, generation-interval mean ∈ {4.7, 5.2, 5.7} d, and event-delay mean ∈ {9, 11, 13} d (27 cells), the behavior-resolved 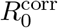 is stable while the scalar bias 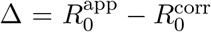 is not even sign-stable (Table 2): Δ ranges over [−0.24, 0.42] and changes sign with the assumed infectious period *v*^−1^. The time-resolved *R*_eff_ wedge and the cross-state confound are qualitatively invariant. We emphasize that the sign instability of Δ is not noise but the scalar-level fingerprint of the non-identifiability (§4.4): a quantity whose very sign flips under innocuous, defensible changes to the assumed natural history cannot be carrying a well-posed “behavioral bias” of *R*_0_. The identified object is the composite *βC*(*t*); its scalar reductions are assumption-dependent by construction.

**Table 2:**
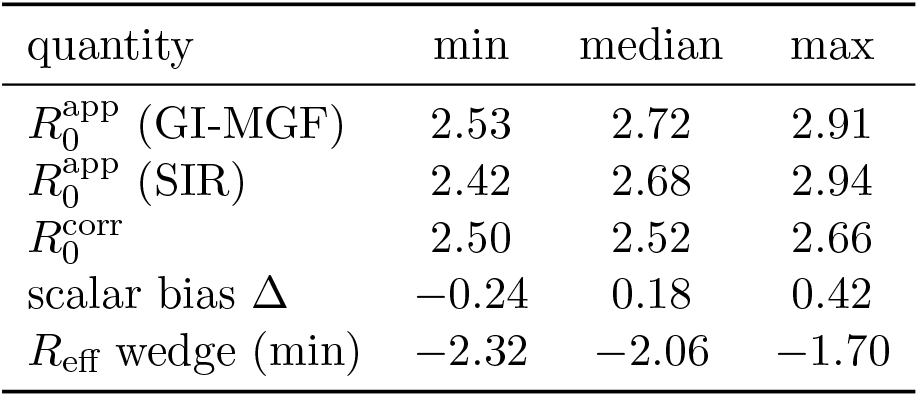
Sensitivity over the 27-cell (*v*, GI, delay) grid. 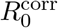 is stable; Δ spans zero.

| quantity | min | median | max |
| --- | --- | --- | --- |
| $R_0^{\text{app}}$ (GI-MGF) | 2.53 | 2.72 | 2.91 |
| $R_0^{\text{app}}$ (SIR) | 2.42 | 2.68 | 2.94 |
| $R_0^{\text{corr}}$ | 2.50 | 2.52 | 2.66 |
| scalar bias $\Delta$ | -0.24 | 0.18 | 0.42 |
| $R_{\text{eff}}$ wedge (min) | -2.32 | -2.06 | -1.70 |

## 5 Discussion

Our result reframes a familiar warning. The concern that *R*_0_ “hides” adaptive behavior is usually voiced as a bias in the scalar estimates too high or too low because behavior contaminates them. We find no support for that version: at baseline the behavior-corrected and naive *R*_0_ nearly coincide, because early transmission precedes the behavioral response. What behavior removes is not a bias in the number but a *non-identifiability* of its decomposition. Two models contact frozen, or contact endogenous to prevalence fit the factual epidemic identically and report the same *R*_0_, yet diverge under any counterfactual that moves the risk trajectory. The apparent *R*_0_ is not wrong; it is incomplete in a way that is invisible in-sample and consequential out-of-sample.

This places the result between the two standard notions of identifiability [19, 20], and it helps to keep them apart. What the trajectory pins down is the composite *βC*(*t*), not its split into a biological rate and a behavioral level: that split is fixed by the normalization *C*(baseline) = 1, not by the data the familiar transmission/susceptibility/reporting trade-off [21, 19]. The scalar reduction 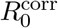 is a milder case of the same thing, stable across parameter inputs but drifting from *≈* 2.5 on the growth window to 3.4–3.6 once the misspecified plateau is included (§4.4) an ordinary *practical*-identifiability symptom [22]. The endogeneity parameter *λ* is different in kind. It is not a parameter of the factual process at all: it enters only the counterfactual map (3), through a term that is identically zero on the factual path (Prop. 1), so a single trajectory run at one operating point, *f* = 1 simply carries no information about it. Recovering it would take variation in *f* itself: distinct variants, waves, or jurisdictions. That makes it a *causal*-identification problem rather than a statistical one the epidemiological counterpart of the Lucas critique [23], in which a relation fitted under one regime need not survive a change of regime. An exogenous-mobility fit is therefore observationally adequate but counterfactually fragile: it silently sets *λ* = 0.

What is deleted is state-dependent, and the response curve explains why. Because *R* saturates, the marginal contact reduction a rise in risk elicits is large only at intermediate risk: a counterfactual intensifying an already-severe epidemic moves behavior along its floor, one that barely produces an epidemic never leaves the flat top, and only intermediate severity rides the steep part of *R* (Fig. 4). The cross-state pattern (§4.6) then explains why the correction is empirically small: the two ingredients of a large one responsiveness and headroom are supplied or withheld together, dense hard-hit jurisdictions tracking risk tightly and cutting mobility to the floor while sparse lightly hit ones kept headroom but never responded. We read this as ecological common cause (density, urbanicity, severity, and stringency driving both axes), though our data identify the coupling, not its source: a crude severity proxy predicts responsiveness (Spearman 0.33) yet not saturation (−0.04), and conditioning on it leaves the association intact (partial −0.32), so a density/urbanicity/stringency-adjusted test (e.g. OxCGRT) is the natural next step. The conclusion holds regardless here the conditions for a large correction do not co-occur and it points to where they would: milder pathogens, graded responses, and relaxation phases that traverse the steep part of *R* from above.

For practice the message is narrow but firm. Description is not at risk reporting *R*_0_ or fitting a wave with exogenous mobility is sound, because *βC*(*t*) is identified whereas any counterfactual that shifts the risk trajectory is, because it turns on the *λ* that an exogenous fit silently sets to zero. The exposure is uneven: attack-rate predictions are *λ*-robust to within a few percent while peak height and timing are not, so surge-capacity planning above all needs the interval. We condense the operational implications below.

**Rules of thumb for practitioners**

1. *Description is safe*. Exogenous-mobility models identify *βC*(*t*); report *R*_0_ as pertaining to its behavioral regime.
2. *Counterfactuals need care*. Any scenario that moves the risk trajectory depends on the unidentified *λ*; exogenous forcing assumes *λ* = 0.
3. *Bound it, or estimate R*. Report the identified set over *λ ∈* [0, 1], or estimate the response (2) to pin the endogenous endpoint.
4. *Know what is exposed*. Attack rate is *λ*-robust; peak height and timing are not surge planning needs the interval.
5. *Screen quickly. λ* matters only when the counterfactual enters the steep part of *R*: greatest at intermediate severity, negligible under saturation.

Several limitations bound these claims. Mobility is a proxy, not contact: *β* absorbs a possibly nonlinear, time-varying mobility-to-contact map, and masking or ventilation go unmeasured. The well-mixed model over-predicts the post-peak decline because once mobility cratered residual transmission moved to household and congregate settings that transit mobility does not capture; we therefore identify *β* from the growth phase only and read the plateau as evidence of the proxy’s limits rather than as signal. Mobility and incidence are jointly determined, and although we use lagged perceived risk the standard identifying assumption we do not claim clean causal identification of *R*. The per-state calibration is coarse, so the confound conclusion is robust while individual span magnitudes are not precise. Finally, the empirical smallness of the deletion is specific to the COVID-19 first wave; the structural results (Prop. 1–2, the regime map) are model-level and carry over, predicting a larger deletion for milder pathogens, graded responses, and relaxation phases. A spatial treatment where the next-generation operator is non-normal and behavior responds to local risk is a natural next step, but our preliminary exploration found the interaction of endogenous local behavior with operator non-normality confounded with the timing of effort and dependent on network structure in ways we could not reduce to a clean law, so we leave it to future work.

## Data Availability

All data produced in the present study are available upon reasonable request to the authors.

## Data and code availability

All data are public: NYC daily counts by event date [11]; US state-level counts [12]; and Google COVID-19 Community Mobility Reports [13], obtained via the archived mirror [14]. All quantities, figures, and tables are reproduced by the analysis code principal scripts `behavioral_r0.py` (apparent/growth-phase *R*_0_), `run_endogenous.py` (response fit, observational equivalence, counter-factuals), `run_regime map.py` and the uncertainty propagation (regime map and Fig. 5), confound check.py (cross-state panel), and `sweep.py` (sensitivity grid). The code and a frozen copy of the derived data will be deposited in a public repository with a citable DOI on acceptance; the archive is available to reviewers on request in the interim.

## Competing interests

The author declares no competing interests. Neither the author nor his institution received any payments or services in the past 36 months from any third party that could be perceived to influence, or give the appearance of potentially influencing, the submitted work.

## Funding

This research received no specific grant from any funding agency in the public, commercial, or not-for-profit sectors. Neither the author nor his institution received, at any time, payment or services from any third party for any aspect of the submitted work.

## A Generation-interval MGF used in 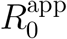

For a renewal process with generation-interval density *g*, the Euler–Lotka relation gives *R*_0_ = 1*/M*_*g*_(−*r*), where 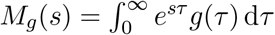 is the moment-generating function and *r* the exponential growth rate [18]. We take *g* gamma with mean *µ* = 5.2 d and standard deviation *σ* = 3.9 d [16], i.e. shape *k* = *µ*^2^*/σ*^2^ *≈* 1.78 and scale *θ* = *σ*^2^*/µ ≈* 2.93 d. The gamma MGF is *M*_*g*_(*s*) = (1 − *θs*)^−*k*^ for *s <* 1*/θ*, so

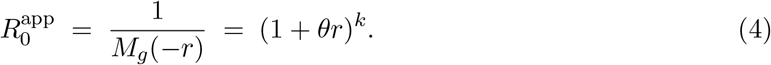

At *r* = 0.258 d^−1^ this gives 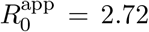 the SIR special case (*g* exponential, *k* = 1, *θ* = 1*/v*) reduces to 1 + *r/v* = 2.68. The 95% interval quoted in the text propagates the regression SE of *r* through this expression.

## Acknowledgments

F.S. extends his gratitude to the Research Center in Pure and Applied Mathematics and the Department of Mathematics at the University of Costa Rica for their support.

## Notes

### Competing Interest Statement

The authors have declared no competing interest.

